# Dihydroartemisinin–piperaquine plus sulfadoxine-pyrimethamine versus either drug alone for intermittent preventive treatment of malaria in pregnancy: a double-blind, randomized, controlled trial

**DOI:** 10.1101/2025.03.24.25324508

**Authors:** Abel Kakuru, Jimmy Kizza, Miriam Aguti, Harriet Adrama, John Ategeka, Peter Olwoch, Miriam Nakalembe, Joaniter I. Nankabirwa, Bishop Opira, Nida Ozarslan, Anju Ranjit, Erin dela Cruz, Tamara D. Clark, Michelle E. Roh, Stephanie L. Gaw, Prasanna Jagannathan, Philip J. Rosenthal, Moses R. Kamya, Grant Dorsey

## Abstract

**Background:** To mitigate adverse consequences of malaria in pregnancy, the World Health Organization recommends intermittent preventive treatment (IPTp) with sulfadoxine-pyrimethamine. However, the effectiveness of IPTp with sulfadoxine-pyrimethamine has been threatened by widespread *P. falciparum* resistance, especially in East and southern Africa. For IPTp, dihydroartemisinin-piperaquine has shown superior antimalarial effects compared to sulfadoxine-pyrimethamine, but sulfadoxine-pyrimethamine has been associated with improved birth outcomes. We hypothesized that a combination of both dihydroartemisinin-piperaquine and sulfadoxine-pyrimethamine would provide superior birth outcomes compared to either drug alone.

**Methods and Findings:** We conducted a double-blinded, randomized, controlled trial of 2757 pregnant women in Uganda, where resistance of malaria parasites to sulfadoxine-pyrimethamine is widespread. Women were randomly assigned (1:1:1) to monthly IPTp with sulfadoxine-pyrimethamine, dihydroartemisinin-piperaquine, or dihydroartemisinin-piperaquine plus sulfadoxine-pyrimethamine. The primary outcome was the risk of a composite adverse birth outcome defined as any of the following: spontaneous abortion, stillbirth, low birthweight (LBW, <2500 gm), preterm delivery (<37 weeks), small-for-gestational age, or neonatal death. Secondary outcomes included specific individual adverse birth outcomes, measures of malaria during pregnancy, and safety.

The risk of a composite adverse birth outcome was lower with sulfadoxine-pyrimethamine compared to dihydroartemisinin-piperaquine (26.4% vs. 30.9%, p=0.04). Combining dihydroartemisinin-piperaquine plus sulfadoxine-pyrimethamine did not reduce the risk of a composite birth outcome compared to dihydroartemisinin-piperaquine (30.0% vs. 30.9%, p=0.70) or sulfadoxine-pyrimethamine (30.0% vs. 26.4%; p=0.10). Considering individual adverse birth outcomes, compared to sulfadoxine-pyrimethamine, dihydroartemisinin-piperaquine was associated with a lower risk of preterm birth (3.1% vs. 5.9%, p=0.007) and a higher risk of small-for-gestational age (22.4% vs. 14.8%, p=0.0006) and low birthweight (6.4% vs. 3.4%, p=0.016) among multigravidae. Combining DP+SP was associated with a higher risk of small-for-gestational age (19.4% vs. 14.8%, p=0.031) and low birthweight (7.1% vs. 3.4%, p=0.0039) among multigravidae compared to sulfadoxine-pyrimethamine. During pregnancy, compared to sulfadoxine-pyrimethamine, dihydroartemisinin-piperaquine was associated with a 94% [88%–97%] reduction in the incidence of symptomatic malaria (0.46 vs. 0.03 episodes per person-year, p<0.0001) and a 97% [95%–98%] reduction in the risk of microscopic parasitaemia (17.7% vs. 0.6%, p<0.0001), but dihydroartemisinin-piperaquine plus sulfadoxine-pyrimethamine was not associated with improved malaria outcomes over dihydroartemisinin-piperaquine alone. There were no significant differences in the incidence of any grade 3-4 adverse events between the treatment arms.

**Conclusions:** Despite the superior antimalarial activity of dihydroartemisinin-piperaquine, sulfadoxine-pyrimethamine was associated with improved birth outcomes. Combining dihydroartemisinin-piperaquine plus sulfadoxine-pyrimethamine for IPTp did not improve birth outcomes compared to either sulfadoxine-pyrimethamine or dihydroartemisinin-piperaquine alone.

**Funding:** National Institute of Allergy and Infectious Diseases, National Institutes of Health (U01AI141308).

**Trial registration:** ClinicalTrials.gov (NCT04336189)

## INTRODUCTION

There were an estimated 36 million pregnancies in African countries with moderate to high malaria transmission in 2023, of which 12.4 million included infection with malaria parasites [1].

Infection with *P. falciparum* during pregnancy is associated with adverse consequences for the mother and fetus, including symptomatic malaria, maternal anemia, fetal loss, preterm birth, low birth weight and neonatal mortality [2]. To mitigate these risks, the World Health Organization has recommended intermittent preventive treatment of malaria in pregnancy (IPTp) with sulfadoxine-pyrimethamine since 1998. This strategy involves the administration of full malaria treatment courses at least one month apart to all at-risk pregnant women starting in the second trimester, and it is currently a component of national malaria policy in 35 African countries [1, 3]. However, the effectiveness of IPTp with sulfadoxine-pyrimethamine has been threatened by widespread *P. falciparum* drug resistance, especially in East and southern Africa, leading to a search for alternative regimens [4].

Dihydroartemisinin-piperaquine has been the most widely studied alternative for IPTp. In a recent meta-analysis of 6 randomized trials, IPTp with dihydroartemisinin-piperaquine was associated with markedly lower risks of both malaria during pregnancy and placental malaria compared to sulfadoxine-pyrimethamine [5]. However, the superior antimalarial activity of dihydroartemisinin-piperaquine did not translate into improved birth outcomes. Rather, compared to sulfadoxine-pyrimethamine, dihydroartemisinin-piperaquine was associated with lower birthweight and a higher risk of infants born small-for-gestational-age.

It remains unclear why IPTp with sulfadoxine-pyrimethamine has been associated with higher birthweight despite the far superior antimalarial activity of dihydroartemisinin-piperaquine. One possible explanation is that sulfadoxine-pyrimethamine, which has antimicrobial activity, improves birthweight through mechanisms independent of its antimalarial activity. This hypothesis is supported by causal mediation analyses showing that effects on birthweight were primarily via non-malarial mechanisms for sulfadoxine-pyrimethamine and antimalarial mechanisms for dihydroartemisinin-piperaquine [5, 6].

We hypothesized that a combination regimen that offered benefits of both dihydroartemisinin-piperaquine and sulfadoxine-pyrimethamine would provide superior birth outcomes compared to either drug alone. The underlying premise for this hypothesis was that dihydroartemisinin-piperaquine would best reduce the risk of malaria-attributable adverse birth outcomes, while sulfadoxine-pyrimethamine would best reduce the risk of adverse birth outcomes attributed to non-malarial mechanisms. To test this hypothesis, we conducted a randomized controlled trial of monthly IPTp with dihydroartemisinin-piperaquine plus sulfadoxine-pyrimethamine compared with either regimen alone in an area of high malaria transmission intensity and widespread resistance to sulfadoxine-pyrimethamine.

## METHODS

### Study design and setting

This was a three-arm, individually randomized, double-blind, controlled trial conducted in Busia District, southeastern Uganda, where malaria transmission is perennial and intense. The study was registered with ClinicalTrials.gov (NCT04336189 https://clinicaltrials.gov/) and was approved by the Makerere University School of Biomedical Sciences Research Ethics Committee (SBS 714), the Uganda National Council for Science and Technology (HS 2746), the Uganda National Drug Authority (CTC 0135/2020), and the University of California San Francisco Human Research Protection Program (19-29105).

### Participants

Study participants were HIV uninfected women with a viable, singleton intrauterine pregnancy at 12-20 weeks gestational age confirmed by ultrasound. Study participants provided written informed consent, agreed to come to a dedicated study clinic for all routine medical care and avoid medication given outside the study clinic, and planned to deliver at a health facility. Women were excluded if they had a history of a serious adverse events from sulfadoxine-pyrimethamine or dihydroartemisinin-piperaquine, active labour, or a history of taking any antimalarial during this pregnancy.

### Randomization and masking

Women were randomly assigned to receive monthly IPTp with sulfadoxine-pyrimethamine, dihydroartemisinin-piperaquine, or dihydroartemisinin-piperaquine plus sulfadoxine-pyrimethamine Randomization was done in a 1:1:1 ratio using permuted blocks of 6 or 9. A computer generated randomization list including consecutive treatment numbers with corresponding random treatment assignments was generated by a study investigator (GD) not involved with the conduct of the field work. Prior to the onset of the study, a set of sequentially numbered, opaque, sealed envelopes containing treatment allocation numbers was prepared. Study pharmacists who were not otherwise involved in the trial were responsible for treatment assignment and preparation of study drugs.

Placebos were used such that all participants received the same number of pills with the same appearance. Study participants, investigators, and study staff involved in the daily care of study participants and assessing study outcomes were blinded to the intervention assigned.

### Study procedures

At enrollment, participants received a long-lasting insecticidal net (LLIN), underwent a standardized examination, and had blood samples collected. Participants received all their medical care at a study clinic open every day.

Each course of sulfadoxine–pyrimethamine consisted of three tablets (500 mg sulfadoxine and 25 mg pyrimethamine [Kamsidar, Kampala Pharmaceutical Industries]) as a single dose. Each course of dihydroartemisinin-piperaquine consisted of three tablets (40 mg dihydroartemisinin and 320 mg piperaquine [Duo-Cotecxin, Holley-Cotec]) once daily for three consecutive days. Study drugs were administered every 4 weeks, starting at 16 or 20 weeks gestation and continued to delivery, up to 40 weeks gestation. Placebos were used such that every 4 weeks participants received either active sulfadoxine–pyrimethamine plus placebo dihydroartemisinin-piperaquine, or placebo sulfadoxine– pyrimethamine plus active dihydroartemisinin-piperaquine, or active sulfadoxine–pyrimethamine plus active dihydroartemisinin-piperaquine. Administration of the first daily dose was directly observed in the clinic, with the second and third doses administered at home. Adherence to study drugs administered at home was assessed by participant recall at their next routine visit.

Routine visits were scheduled every 4 weeks for administration of study drugs, a clinical examination, and a blood draw for detection of malaria parasites by microscopy and quantitative PCR (qPCR). Complete blood counts were performed at 20, 28, and 36 weeks gestation. Participants were encouraged to come to the clinic any time they required medical care. At any visit, participants with a history of fever in the past 24 hours or a tympanic temperature ≥38.0°C had a thick blood smear assessed for malaria parasites. If the smear was positive, the participant was diagnosed with symptomatic malaria and treated with artemether-lumefantrine. Participants with asymptomatic parasitaemia detected at the time of routine visits were not provided additional antimalarial therapy beyond their assigned IPTp drugs in accordance with local guidelines.

Study participants were encouraged to deliver at a health facility. Those who delivered at home were visited by study staff shortly after delivery and encouraged to come to the study clinic. At delivery, a standardized assessment was conducted, including birthweight, evaluation for congenital anomalies, and collection of placental tissue and maternal, placental, and cord blood. Participants were followed for 4 weeks post-partum. Adverse events were assessed and graded at every clinic visit according to standardized criteria [7]. For the first 300 participants enrolled, electrocardiograms were done at 20, 28, and 36 gestation weeks just before the first dose and 2-6 hours after the third dose of study drugs.

Blood smears were stained with 2% Giemsa and considered negative when 100 high power fields did not reveal asexual parasites. All slides were read by a second microscopist, and a third reviewer settled any discrepant readings. A highly sensitive *P. falciparum* qPCR assay with a lower limit of detection of 1 parasite/µL was performed on blood collected at enrollment, routine visits, and delivery [8].

### Outcomes

The primary outcome was the risk of a composite adverse birth outcome defined as any of the following: spontaneous abortion (fetal loss at <28 weeks gestational age), stillbirth (infants born deceased at >28 weeks gestational age), and among live births: low birthweight (LBW, <2500 gm), preterm delivery (<37 weeks), small-for-gestational age (birthweight <10^th^ percentile for gestational age by INTERGROWTH-21^st^ standards) [9], or neonatal death (within the first 28 days of life).

Secondary outcomes included individual components of the primary outcome, birthweight, gestational age at birth, birthweight-for-gestational age z-score [9], and gestational weight gain.

Malaria related outcomes measured following the initiation of study drugs included the incidence of symptomatic malaria; prevalence of parasitaemia by microscopy or qPCR at routine visits; prevalence of any (Hb <11 gm/dL) or severe (Hb <8 gm/dL) anemia at 20, 28, or 36 gestational weeks; detection of malaria parasites by microscopy or qPCR at birth from maternal, placental, or cord blood; and measures of placental malaria by histopathology, including detection of parasites (active infection), malaria pigment in ≥30% of high powered fields (high grade past infection) [10], or any parasites or malaria pigment (any active or past infection). Measures of tolerability and safety included the incidence of vomiting, common adverse events of any severity, and serious adverse events following administration of study drugs.

### Statistical analysis

We assumed a risk of composite adverse birth outcome of 22.6% in the sulfadoxine-pyrimethamine arm and 25.1% in the dihydroartemisinin-piperaquine arm based on prior data [11]. A sample size of 2757 (assuming 15% loss to follow-up) was required to achieve 80% power (2-sided alpha = 0.05) to detect a ≥25% reduction with dihydroartemisinin-piperaquine plus sulfadoxine-pyrimethamine compared to the other arms. Statistical analyses were conducted using Stata (version 18). All analyses used a modified intention-to-treat approach and included all subjects with evaluable outcomes. Comparisons of the primary outcome and dichotomous secondary outcomes were made using log-binomial regression to obtain risk ratios (RRs). Comparisons of dichotomous secondary outcomes with repeated measures (parasite prevalence and anemia during pregnancy) were performed using generalized estimating equations with a log-binomial model and robust standard errors. Comparisons of incidence measures were performed using negative binomial regression to obtain incidence rate ratios (IRRs). Comparisons of continuous outcomes were performed using linear regression to obtain mean differences. The number needed to treat to avert one event was calculated as the inverse of the incidence rate difference per 21.4 weeks (the average duration of follow-up following study drug initiation). Subgroup analyses by gravidity were pre-specified, and differences were tested using two-way interaction terms between treatment arm and gravidity subgroup (Pinteraction). Statistical significance was defined as a two-sided p value of <0.05.

## RESULTS

### Study participants and follow-up

Between December 28, 2020, and December 18, 2023, 3063 women were screened, of whom 2757 were enrolled and underwent randomization (Figure 1). Baseline characteristics were similar between treatment arms (Table 1). The mean age at enrollment was 24.5 years (standard deviation 6.2 years), 1533 (55.6%) of 2757 participants were enrolled between 12-16 gestational weeks, 726 (26.3%) were primigravidae, and 1360 (49.3%) reported owning an LLIN at the time of enrollment (all participants were given an LLIN after enrollment). Parasite prevalence was 38.0% (1047/2757) by microscopy and 70.3% (1937/2757) by microscopy or qPCR. In a random subset of 200 enrollment samples with parasite densities >100/µL, 198 (99%) had five *P. falciparum* dihydrofolate reductase (PfDHFR; N51I, C59R, and S108N) and dihydropteroate synthetase (PfDHPS; A437G and K540E) mutations associated with resistance to sulfadoxine-pyrimethamine and known to be common in Uganda.[12] Considering mutations associated with higher-level resistance, 41 (20.5%) of 200 had the PfDHFR I164L and four (2%) had the PfDHPS A581G mutation. A total of 2706 (98.2%) participants enrolled received at least one dose of study drugs, and 2538 (92.1%) were followed through delivery (Figure 1).

**Figure 1.**
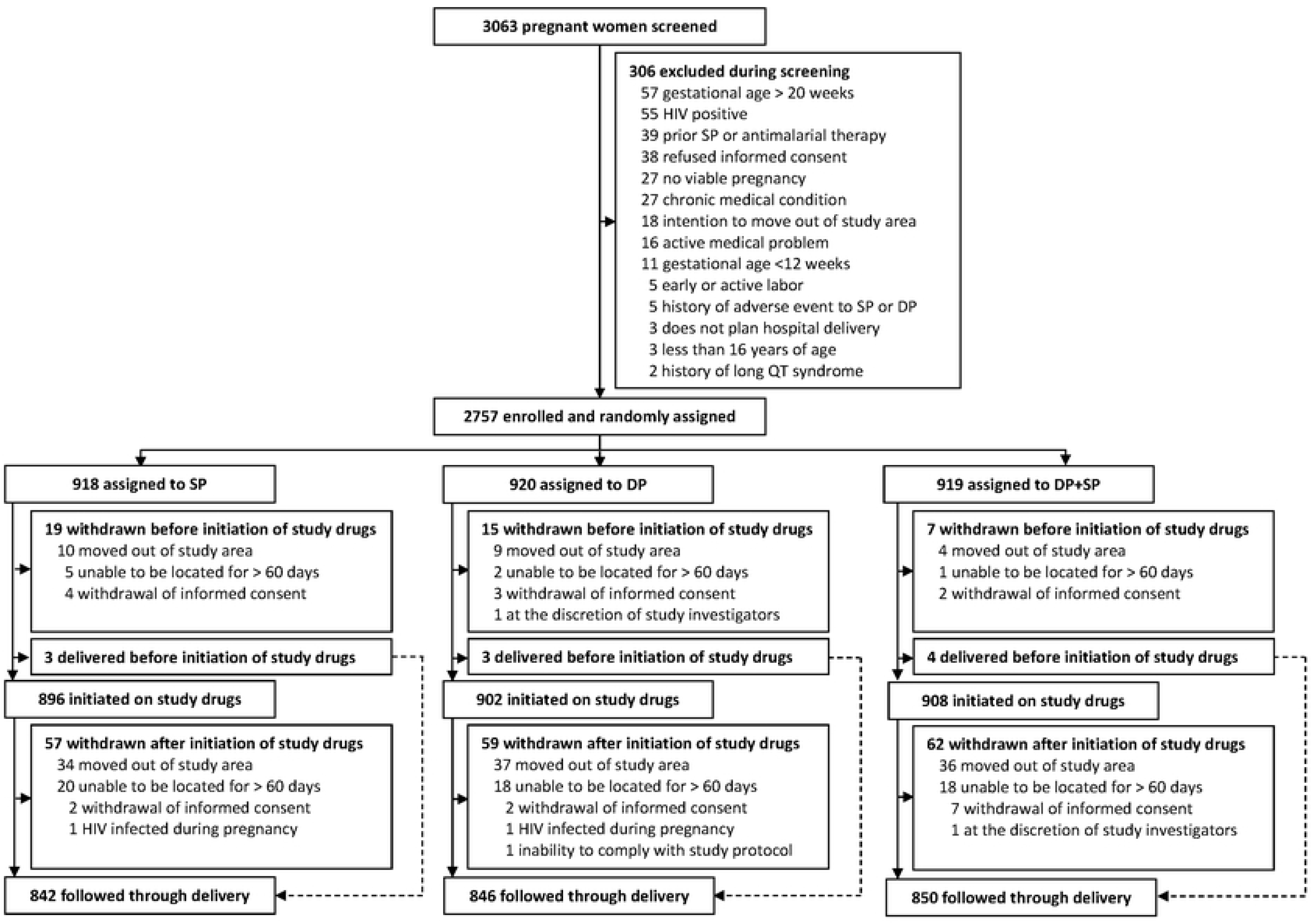
Trial profile

**Table 1.** Baseline characteristics of enrolled study participants.

| Characteristic | Treatment arm |  |  |
| --- | --- | --- | --- |
|  | SP<sup>1</sup><br>(n=918) | DP<sup>1</sup><br>(n=920) | DP+SP<sup>1</sup><br>(n=919) |
| Age in years, mean (SD) | 24.5 (6.2) | 24.7 (6.3) | 24.2 (6.0) |
| Gestational age in weeks, mean (SD) | 15.8 (2.6) | 15.7 (2.5) | 16.0 (2.6) |
| Gestational age categories, n (%) |  |  |  |
| 12-16 weeks | 522 (56.9%) | 530 (57.6%) | 481 (52.3%) |
| >16-20 weeks | 396 (43.1%) | 390 (42.4%) | 438 (47.7%) |
| Gravidity, n (%) |  |  |  |
| Primigravidae | 230 (25.1%) | 234 (25.4%) | 262 (28.5%) |
| Multigravidae | 688 (74.9%) | 686 (74.6%) | 657 (71.5%) |
| Bednet ownership, n (%) |  |  |  |
| None | 423 (46.1%) | 434 (47.2%) | 415 (45.2%) |
| Untreated net | 43 (4.7%) | 40 (4.4%) | 42 (4.6%) |
| Long-lasting insecticide-treated net | 452 (49.2%) | 446 (48.5%) | 462 (50.3%) |
| Household wealth index, n (%) |  |  |  |
| Lowest tertile | 308 (33.9%) | 294 (32.3%) | 295 (32.4%) |
| Middle tertile | 293 (32.3%) | 319 (35.1%) | 309 (33.9%) |
| Highest tertile | 307 (33.8%) | 296 (32.6%) | 307 (33.7%) |
| Highest level of education, n (%) |  |  |  |
| None | 44 (4.8%) | 31 (3.4%) | 36 (3.9%) |
| Primary school | 592 (64.5%) | 593 (64.5%) | 600 (65.3%) |
| Secondary school | 263 (28.7%) | 276 (30.0%) | 264 (28.7%) |
| Higher | 19 (2.1%) | 20 (2.2%) | 19 (2.1%) |
| Weight in kg, mean (SD) | 57.3 (9.4) | 57.6 (9.8) | 57.1 (8.9) |
| Height in cm, mean (SD) | 159 (6) | 159 (7) | 159 (6) |
| Maternal MUAC, mean (SD) | 26 (4) | 26 (3) | 26 (3) |
| Mean hemoglobin g/dL (SD) | 11.6 (1.3) | 11.7 (1.3) | 11.5 (1.4) |
| Detection of malaria parasites by microscopy, n (%) | 332 (36.2%) | 347 (37.7%) | 368 (40.0%) |
| Detection of malaria parasites by microscopy or qPCR, n (%) | 643 (70.1%) | 637 (69.2%) | 657 (71.5%) |
| Prevalence of markers of antifolate resistance, n (%)<sup>2</sup> | N=72 | N=65 | N=63 |
| *dhfr/dhps* quintuple mutant<sup>3</sup> | 72 (100%) | 64 (98.5%) | 62 (98.4%) |
| *dhfr* I164L | 14 (19.4%) | 19 (29.2%) | 8 (12.7%) |
| *dhps* A581G | 1 (1.4%) | 2 (3.1%) | 1 (1.6%) |

<sup>1</sup> SP = sulfadoxine-pyrimethamine, DP = dihydroartemisinin-piperaquine
<sup>2</sup> Random subset of 200 samples with parasite density > 100/μL; mixed genotypes were categorized as mutant.
<sup>3</sup> *dhfr* N51I+C59R+S108N; *dhps* A437G+K540E

### Efficacy outcomes

IPTp with dihydroartemisinin-piperaquine was associated with a higher risk of the composite adverse birth outcome compared to sulfadoxine-pyrimethamine (261 [30.9%] of 846 vs. 222 [26.4%] of 842; RR 1.17 [95% CI 1.01-1.36], p=0ꞏ042) (Figure 2A). Similarly, IPTp with dihydroartemisinin-piperaquine plus sulfadoxine-pyrimethamine was associated with a higher risk of the composite adverse birth outcome compared to sulfadoxine-pyrimethamine alone, although this difference did not reach statistical significance (255 [30.0%] of 850 vs. 222 [26.4%] of 842; RR 1.14 [95% CI 0.98- 1.33], p=0.097) (Figure 2B). IPTp with dihydroartemisinin-piperaquine plus sulfadoxine-pyrimethamine was associated with a similar risk of the composite adverse birth outcome compared to dihydroartemisinin-piperaquine alone (255 [30.0%] of 850 vs. 261 [30.9%] of 846; RR 0.97 [95% CI 0.84-1.12], p=0.70) (Figure 2C). In summary, dihydroartemisinin-piperaquine was associated with a higher risk of the adverse birth outcome compared to sulfadoxine-pyrimethamine, and combining the two regimens did not lower this risk.

**Figure 2.**
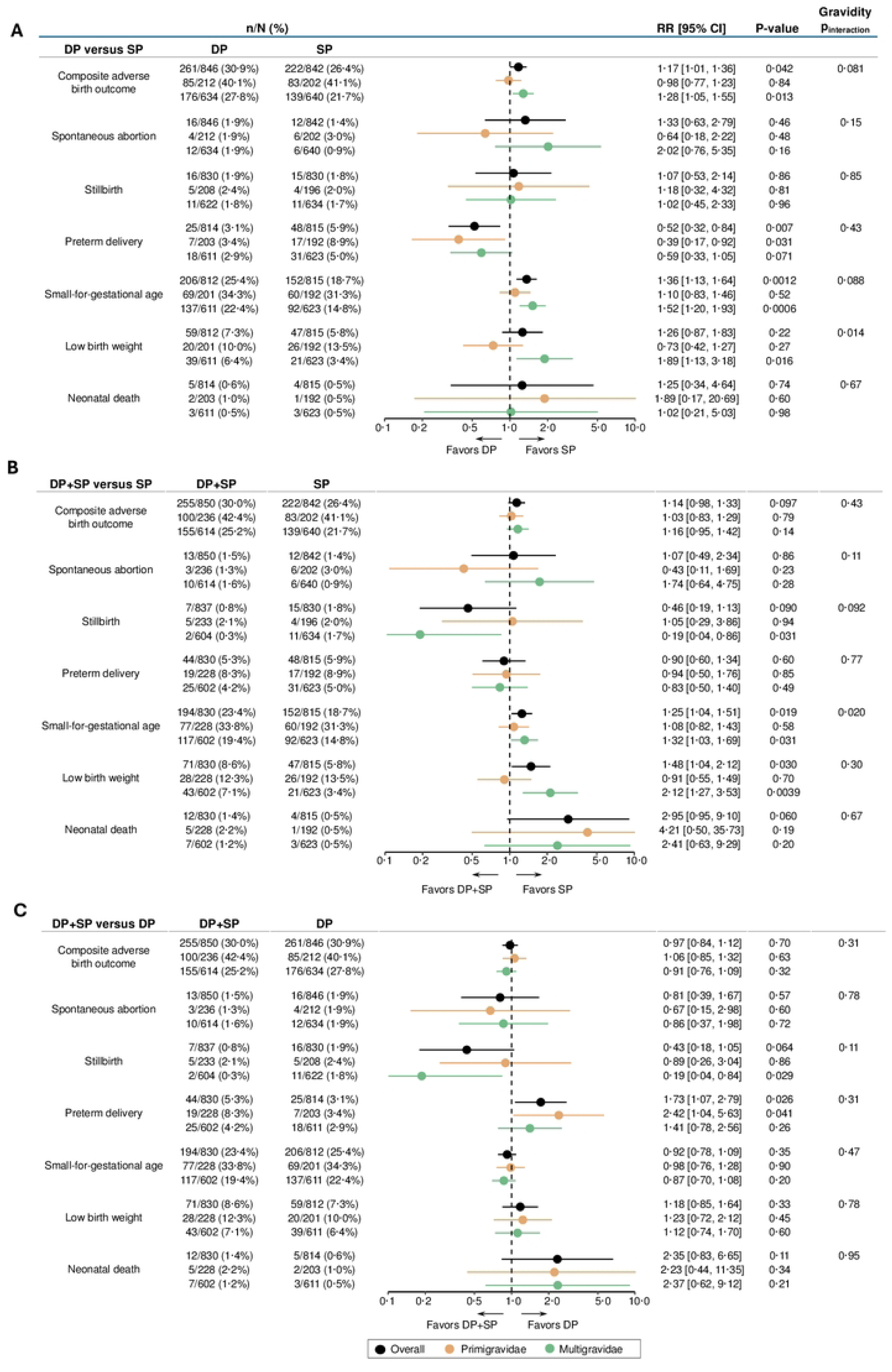
Primary endpoint and individual adverse birth outcomes including stratification by gravidity. All pair-wise comparisons included in sub-figures A-C. DP = dihydroartemisinin-piperaquine, SP = sulfadoxine-pyrimethamine, and RR = relative risk. Pinteraction denotes p-values from two-way interaction terms evaluating differences in treatment effects between gravidity subgroups.

When stratified by gravidity, the higher risk of a composite adverse birth outcome with dihydroartemisinin-piperaquine compared to sulfadoxine-pyrimethamine was present among multigravidae (176 [27.8%] of 634 vs. 139 [21.7%] of 640, RR 1ꞏ28 [95% CI 1ꞏ05-1ꞏ55], p=0ꞏ013) but not primigravidae (85 of [40ꞏ1%] of 212 vs. 83 [41.1%] of 202; RR 0.98 [95% CI 0.77-1.23], p=0.84). Considering individual adverse birth outcomes stratified by gravidity, among primigravidae, dihydroartemisinin-piperaquine was associated with a lower risk of preterm delivery compared to sulfadoxine-pyrimethamine (7 [3.4%] of 203 vs. 17 [8.9%] of 192; RR 0.39 [95% CI 0.17-0.92], p=0.031), with no significant differences for the other adverse birth outcomes. In contrast, among multigravidae, dihydroartemisinin-piperaquine was associated with a higher risk of small-for-gestational age (137 [22.4%] of 611 vs. 92 [14.8%] of 623; RR 1.52 [95% CI 1.20-1.93], p=0.0006) and LBW (39 [6.4%] of 611 vs. 21 [3.4%] of 623; RR 1.89 [95% CI 1.13-3.18], p=0.016) compared to sulfadoxine-pyrimethamine, with no significant differences for the other adverse birth outcomes (Figure 2A). Among primigravidae, there were no significant differences in the risk of individual adverse birth outcomes between the dihydroartemisinin-piperaquine plus sulfadoxine-pyrimethamine and sulfadoxine-pyrimethamine alone arms. Among multigravidae, dihydroartemisinin-piperaquine plus sulfadoxine-pyrimethamine was associated with a higher risk of small-for-gestational age (117 [19.4%] of 602 vs. 21 [14.8%] of 623; RR 1.32 [95% CI 1.03-1.69], p=0.031) and LBW (43 [7.1%] of 602 vs. 21 [3.4%] of 623, RR 2.12 [95% CI 1.27-3.53], p=0.0039) but a lower risk of stillbirth (2 [0.3%] of 604 vs. 11 [1.7%] of 634; RR 0.19 [95% CI 0.04-0.86], p=0.031) compared to sulfadoxine-pyrimethamine alone (Figure 2B). There were no significant differences in the risk of individual adverse birth outcomes between the dihydroartemisinin-piperaquine plus sulfadoxine-pyrimethamine and dihydroartemisinin-piperaquine alone arms, with the exception of a higher risk of preterm delivery (19 [8.3%] of 228 vs. 7 [3.4%] of 192; RR 2.42 [95% CI 1.04-5.63], p=0.041) among primigravidae and a lower risk of stillbirth (2 [0.3%] of 604 vs. 11 [1.8%] of 622; RR 0.19 [95% CI 0.04-0.84], p=0.029) among multigravidae and in the dihydroartemisinin-piperaquine plus sulfadoxine-pyrimethamine arm (Figure 2C).

Considering continuous outcomes, dihydroartemisinin-piperaquine was associated with a lower mean birthweight (3057 vs. 3123 gm; MD -66 [95% CI -112, -20], p=0.0052), lower birthweight-for-gestational age z-scores (-0.58 vs. -0.37; MD -0.21 [95% CI -0.31, -0.12], p<0.0001), and lower gestational weight gain (220 vs. 256 gm/week; MD -36 [95% CI -49, -22], p<0.0001) compared to sulfadoxine-pyrimethamine, with differences greater among multigravidae compared to primigravidae (Figure 3A). Similarly, dihydroartemisinin-piperaquine plus sulfadoxine-pyrimethamine was associated with a lower mean birthweight (3068 vs. 3123 gm; MD -55 [95% CI -103, -7], p=0.025), lower birthweight-for-gestational age z-scores (-0.49 vs. -0.37; MD -0.12 [95% CI -0.22, -0.02], p=0.015), and lower gestational weight gain (236 vs. 256 gm/week; MD -21 [95% CI -34, -7], p=0.0021) compared to sulfadoxine-pyrimethamine alone, with differences greater among multigravidae compared to primigravidae (Figure 3B). There were no significant differences in gestational age at delivery between any of the treatment arms or other continuous outcomes when comparing dihydroartemisinin-piperaquine plus sulfadoxine-pyrimethamine to dihydroartemisinin-piperaquine alone with the exception of higher gestational weight gain (240 vs. 222 gm/week; MD 18 [95% CI 2, 33], p=0.026) among multigravidae in the dihydroartemisinin-piperaquine plus sulfadoxine-pyrimethamine arm (Figure 3C).

**Figure 3.**
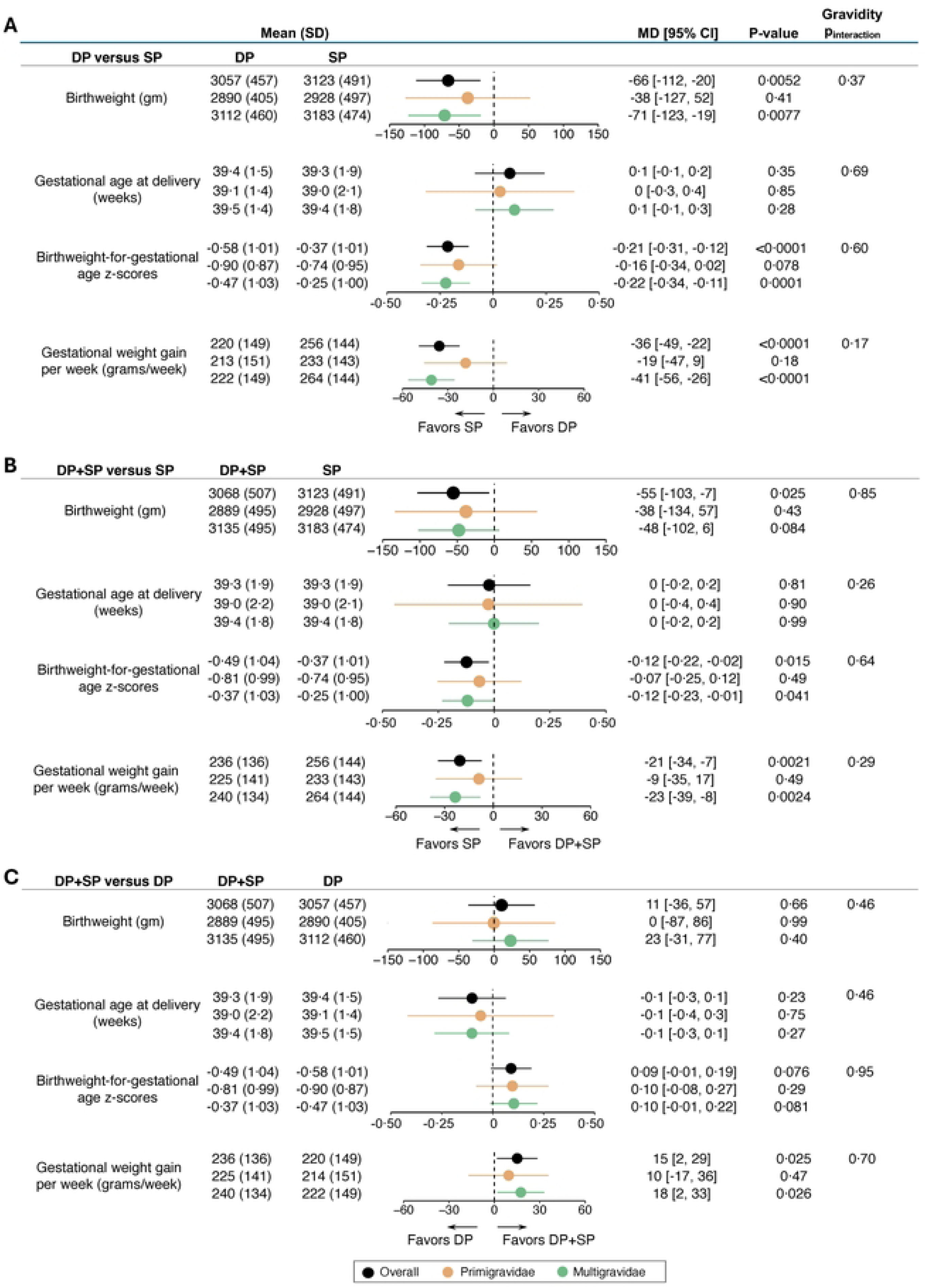
Continuous maternal and birth outcomes including stratification by gravidity. All pair-wise comparisons included in sub-figures A-C. DP = dihydroartemisinin-piperaquine, SP = sulfadoxine-pyrimethamine, and MD = mean difference. Pinteraction denotes p-values from two-way interaction terms evaluating differences in treatment effects between gravidity subgroups.

During pregnancy, dihydroartemisinin-piperaquine was associated with a 94% reduction in the incidence of symptomatic malaria (0.03 vs. 0.46 episodes per person year; IRR 0.06 [95% CI 0.03- 0.12], p<0.0001), 97% reduction in the risk of microscopic parasitaemia (0.6% vs. 17.7%; RR 0.03 [95% CI 0.02-0.05], p<0.0001), 15% reduction in the risk of any anemia (36.7% vs. 43.2%; RR 0.85 [95% CI 0.77-0.94], p=0.0012), and marked reductions in risks of various measures of malaria at delivery compared to sulfadoxine-pyrimethamine (Table 2). As expected, the burden of malaria was higher in primigravidae compared to multigravidae, resulting in greater absolute malaria-related benefits of dihydroartemisinin-piperaquine over sulfadoxine-pyrimethamine among primigravidae compared to multigravidae (Table 3). For example, the number needed to treat with dihydroartemisinin-piperaquine vs. sulfadoxine-pyrimethamine to avert one episode of symptomatic malaria was approximately 2 among primigravidae compared to 11 among multigravidae. Similarly, dihydroartemisinin-piperaquine plus sulfadoxine-pyrimethamine was associated with a 95% reduction in the incidence of symptomatic malaria (0.02 vs. 0.46 episodes per person year; IRR 0.05 [95% CI 0.03-0.10], p<0.0001), 92% reduction in the risk of microscopic parasitaemia (1.4% vs. 17.7%; RR 0.08 [95% CI 0.06-0.10], p<0.0001), 10% reduction in the risk of any anemia (39.5% vs. 43.2%; RR 0.90 [95% CI 0.82-0.99], p=0.034), and marked reductions in risks of various measures of malaria at delivery compared to sulfadoxine-pyrimethamine alone (Table 2). Compared to dihydroartemisinin-piperaquine alone, dihydroartemisinin-piperaquine plus sulfadoxine-pyrimethamine did not improve malaria-related outcomes and was associated with higher risks of parasitaemia (1.4% vs. 0.6%; RR 2.35 [95% CI 1.34-0.10], p=0.0026) during pregnancy and any evidence of placental malaria by histopathology (50.1% vs. 40.8%; RR 1.23 [95% CI 1.10-1.37], p=0.0002) (Table 2). In summary, dihydroartemisinin-piperaquine was markedly superior to sulfadoxine-pyrimethamine in regard to malaria outcomes during pregnancy, but combining the two regimens did not offer benefits against malaria outcomes compared to dihydroartemisinin-piperaquine alone.

**Table 2.**
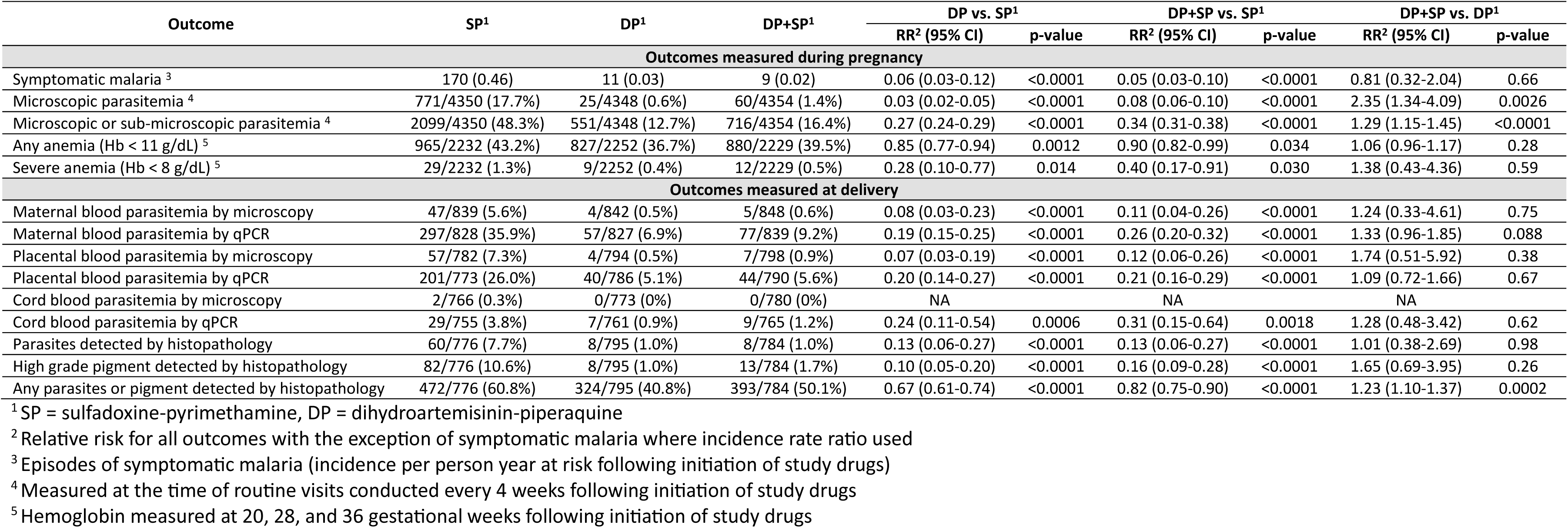
Secondary efficacy outcomes.

| Outcome | SP<sup>1</sup> | DP<sup>1</sup> | DP+SP<sup>1</sup> | DP vs. SP<sup>1</sup> |  | DP+SP vs. SP<sup>1</sup> |  | DP+SP vs. DP<sup>1</sup> |  |
| --- | --- | --- | --- | --- | --- | --- | --- | --- | --- |
|  |  |  |  | RR<sup>2</sup> (95% CI) | p-value | RR<sup>2</sup> (95% CI) | p-value | RR<sup>2</sup> (95% CI) | p-value |
| Outcomes measured during pregnancy |  |  |  |  |  |  |  |  |  |
| Symptomatic malaria <sup>3</sup> | 170 (0.46) | 11 (0.03) | 9 (0.02) | 0.06 (0.03-0.12) | <0.0001 | 0.05 (0.03-0.10) | <0.0001 | 0.81 (0.32-2.04) | 0.66 |
| Microscopic parasitemia <sup>4</sup> | 771/4350 (17.7%) | 25/4348 (0.6%) | 60/4354 (1.4%) | 0.03 (0.02-0.05) | <0.0001 | 0.08 (0.06-0.10) | <0.0001 | 2.35 (1.34-4.09) | 0.0026 |
| Microscopic or sub-microscopic parasitemia <sup>4</sup> | 2099/4350 (48.3%) | 551/4348 (12.7%) | 716/4354 (16.4%) | 0.27 (0.24-0.29) | <0.0001 | 0.34 (0.31-0.38) | <0.0001 | 1.29 (1.15-1.45) | <0.0001 |
| Any anemia (Hb < 11 g/dL) <sup>5</sup> | 965/2232 (43.2%) | 827/2252 (36.7%) | 880/2229 (39.5%) | 0.85 (0.77-0.94) | 0.0012 | 0.90 (0.82-0.99) | 0.034 | 1.06 (0.96-1.17) | 0.28 |
| Severe anemia (Hb < 8 g/dL) <sup>5</sup> | 29/2232 (1.3%) | 9/2252 (0.4%) | 12/2229 (0.5%) | 0.28 (0.10-0.77) | 0.014 | 0.40 (0.17-0.91) | 0.030 | 1.38 (0.43-4.36) | 0.59 |
| Outcomes measured at delivery |  |  |  |  |  |  |  |  |  |
| Maternal blood parasitemia by microscopy | 47/839 (5.6%) | 4/842 (0.5%) | 5/848 (0.6%) | 0.08 (0.03-0.23) | <0.0001 | 0.11 (0.04-0.26) | <0.0001 | 1.24 (0.33-4.61) | 0.75 |
| Maternal blood parasitemia by qPCR | 297/828 (35.9%) | 57/827 (6.9%) | 77/839 (9.2%) | 0.19 (0.15-0.25) | <0.0001 | 0.26 (0.20-0.32) | <0.0001 | 1.33 (0.96-1.85) | 0.088 |
| Placental blood parasitemia by microscopy | 57/782 (7.3%) | 4/794 (0.5%) | 7/798 (0.9%) | 0.07 (0.03-0.19) | <0.0001 | 0.12 (0.06-0.26) | <0.0001 | 1.74 (0.51-5.92) | 0.38 |
| Placental blood parasitemia by qPCR | 201/773 (26.0%) | 40/786 (5.1%) | 44/790 (5.6%) | 0.20 (0.14-0.27) | <0.0001 | 0.21 (0.16-0.29) | <0.0001 | 1.09 (0.72-1.66) | 0.67 |
| Cord blood parasitemia by microscopy | 2/766 (0.3%) | 0/773 (0%) | 0/780 (0%) | NA |  | NA |  | NA |  |
| Cord blood parasitemia by qPCR | 29/755 (3.8%) | 7/761 (0.9%) | 9/765 (1.2%) | 0.24 (0.11-0.54) | 0.0006 | 0.31 (0.15-0.64) | 0.0018 | 1.28 (0.48-3.42) | 0.62 |
| Parasites detected by histopathology | 60/776 (7.7%) | 8/795 (1.0%) | 8/784 (1.0%) | 0.13 (0.06-0.27) | <0.0001 | 0.13 (0.06-0.27) | <0.0001 | 1.01 (0.38-2.69) | 0.98 |
| High grade pigment detected by histopathology | 82/776 (10.6%) | 8/795 (1.0%) | 13/784 (1.7%) | 0.10 (0.05-0.20) | <0.0001 | 0.16 (0.09-0.28) | <0.0001 | 1.65 (0.69-3.95) | 0.26 |
| Any parasites or pigment detected by histopathology | 472/776 (60.8%) | 324/795 (40.8%) | 393/784 (50.1%) | 0.67 (0.61-0.74) | <0.0001 | 0.82 (0.75-0.90) | <0.0001 | 1.23 (1.10-1.37) | 0.0002 |

<sup>1</sup> SP = sulfadoxine-pyrimethamine, DP = dihydroartemisinin-piperaquine
<sup>2</sup> Relative risk for all outcomes with the exception of symptomatic malaria where incidence rate ratio used
<sup>3</sup> Episodes of symptomatic malaria (incidence per person year at risk following initiation of study drugs)
<sup>4</sup> Measured at the time of routine visits conducted every 4 weeks following initiation of study drugs
<sup>5</sup> Hemoglobin measured at 20, 28, and 36 gestational weeks following initiation of study drugs

**Table 3.** Secondary efficacy outcomes stratified by gravidity.

| Outcome | SP<sup>1</sup> | DP<sup>1</sup> | DP+SP<sup>1</sup> | DP vs. SP<sup>1</sup> |  | DP+SP vs. SP<sup>1</sup> |  | DP+SP vs. DP<sup>1</sup> |  |
| --- | --- | --- | --- | --- | --- | --- | --- | --- | --- |
|  |  |  |  | RR<sup>2</sup> (95% CI) | p-value | RR<sup>2</sup> (95% CI) | p-value | RR<sup>2</sup> (95% CI) | p-value |
| Primigravidae |  |  |  |  |  |  |  |  |  |
| Symptomatic malaria <sup>3</sup> | 99 (1.15) | 2 (0.02) | 4 (0.04) | 0.02 (0.005-0.08) | <0.0001 | 0.03 (0.01-0.09) | <0.0001 | 1.79 (0.26-12.4) | 0.56 |
| Microscopic parasitemia <sup>4</sup> | 360/1004 (35.9%) | 8/1044 (0.8%) | 21/1179 (1.8%) | 0.02 (0.01-0.04) | <0.0001 | 0.05 (0.03-0.08) | <0.0001 | 2.35 (0.97-5.68) | 0.057 |
| Microscopic or sub-microscopic parasitemia <sup>4</sup> | 641/1004 (63.8%) | 161/1044 (15.4%) | 236/1179 (20.0%) | 0.25 (0.21-0.29) | <0.0001 | 0.32 (0.27-0.37) | <0.0001 | 1.30 (1.06-1.59) | 0.012 |
| Any anemia (Hb < 11 g/dL) <sup>5</sup> | 300/511 (58.7%) | 227/542 (41.9%) | 255/596 (42.8%) | 0.70 (0.60-0.82) | <0.0001 | 0.73 (0.62-0.85) | 0.0001 | 1.04 (0.87-1.25) | 0.66 |
| Severe anemia (Hb < 8 g/dL) <sup>5</sup> | 14/511 (2.7%) | 0/542 (0%) | 3/596 (0.5%) | NA |  | 0.16 (0.03-0.74) | 0.020 | NA |  |
| Maternal blood parasitemia by microscopy | 23/201 (11.4%) | 1/212 (0.5%) | 3/236 (1.3%) | 0.04 (0.01-0.30) | 0.0017 | 0.11 (0.03-0.36) | 0.0003 | 2.69 (0.28-25.7) | 0.39 |
| Maternal blood parasitemia by qPCR | 103/198 (52.0%) | 14/210 (6.7%) | 24/233 (10.3%) | 0.13 (0.08-0.22) | <0.0001 | 0.20 (0.13-0.30) | <0.0001 | 1.55 (0.82-2.91) | 0.18 |
| Placental blood parasitemia by microscopy | 29/183 (15.9%) | 1/194 (0.5%) | 4/220 (1.8%) | 0.03 (0.004-0.24) | 0.0007 | 0.11 (0.04-0.32) | <0.0001 | 3.53 (0.40-31.3) | 0.26 |
| Placental blood parasitemia by qPCR | 75/182 (41.2%) | 7/194 (3.6%) | 15/222 (6.8%) | 0.09 (0.04-0.18) | <0.0001 | 0.16 (0.10-0.28) | <0.0001 | 1.87 (0.78-4.50) | 0.16 |
| Cord blood parasitemia by microscopy | 1/181 (0.6%) | 0/188 (0%) | 0/215 (0%) | NA |  | NA |  | NA |  |
| Cord blood parasitemia by qPCR | 10/177 (5.7%) | 1/186 (0.5%) | 5/211 (2.4%) | 0.10 (0.01-0.74) | 0.024 | 0.42 (0.15-1.20) | 0.11 | 4.41 (0.52-37.4) | 0.17 |
| Parasites detected by histopathology | 33/177 (18.6%) | 2/196 (1.0%) | 5/218 (2.3%) | 0.05 (0.01-0.22) | 0.0001 | 0.12 (0.05-0.31) | <0.0001 | 2.25 (0.44-11.5) | 0.33 |
| High grade pigment detected by histopathology | 55/177 (31.1%) | 7/196 (3.6%) | 8/218 (3.7%) | 0.11 (0.05-0.25) | <0.0001 | 0.12 (0.06-0.24) | <0.0001 | 1.03 (0.38-2.78) | 0.96 |
| Any parasites or pigment detected by histopathology | 156/177 (88.1%) | 145/196 (74.0%) | 167/218 (76.6%) | 0.84 (0.76-0.93) | 0.0005 | 0.87 (0.79-0.93) | 0.0026 | 1.03 (0.93-1.16) | 0.54 |
| Multigravidae |  |  |  |  |  |  |  |  |  |
| Symptomatic malaria <sup>2</sup> | 71 (0.25) | 9 (0.03) | 5 (0.02) | 0.13 (0.06-0.26) | <0.0001 | 0.07 (0.03-0.18) | <0.0001 | 0.58 (0.19-1.73) | 0.33 |
| Microscopic parasitemia <sup>4</sup> | 411/3346 (12.3%) | 17/3304 (0.5%) | 39/3175 (1.2%) | 0.04 (0.02-0.08) | <0.0001 | 0.10 (0.07-0.15) | <0.0001 | 2.28 (1.12-4.63) | 0.023 |
| Microscopic or sub-microscopic parasitemia <sup>4</sup> | 1458/3346 (43.6%) | 390/3304 (11.8%) | 480/3175 (15.1%) | 0.27 (0.24-0.31) | <0.0001 | 0.35 (0.31-0.39) | <0.0001 | 1.28 (1.11-1.47) | 0.0007 |
| Any anemia (Hb < 11 g/dL) <sup>5</sup> | 665/1721 (38.6%) | 600/1710 (35.1%) | 625/1633 (38.3%) | 0.93 (0.82-1.04) | 0.20 | 0.98 (0.87-1.10) | 0.74 | 1.06 (0.94-1.20) | 0.35 |
| Severe anemia (Hb < 8 g/dL) <sup>5</sup> | 15/1721 (0.9%) | 9/1710 (0.5%) | 9/1633 (0.6%) | 0.60 (0.19-1.87) | 0.37 | 0.66 (0.22-1.93) | 0.45 | 1.10 (0.32-3.73) | 0.88 |
| Maternal blood parasitemia by microscopy | 24/638 (3.8%) | 3/630 (0.5%) | 2/612 (0.3%) | 0.13 (0.04-0.42) | 0.0007 | 0.09 (0.02-0.37) | 0.0009 | 0.69 (0.12-4.09) | 0.68 |
| Maternal blood parasitemia by qPCR | 194/630 (30.8%) | 43/617 (7.0%) | 53/606 (8.8%) | 0.23 (0.17-0.31) | <0.0001 | 0.28 (0.21-0.38) | <0.0001 | 1.25 (0.85-1.85) | 0.25 |
| Placental blood parasitemia by microscopy | 28/599 (4.7%) | 3/600 (0.5%) | 3/578 (0.5%) | 0.11 (0.03-0.35) | 0.0002 | 0.11 (0.03-0.36) | 0.0003 | 1.04 (0.21-5.12) | 0.96 |
| Placental blood parasitemia by qPCR | 126/591 (21.3%) | 33/592 (5.6%) | 29/568 (5.1%) | 0.26 (0.18-0.38) | <0.0001 | 0.24 (0.16-0.35) | <0.0001 | 0.92 (0.56-1.49) | 0.72 |
| Cord blood parasitemia by microscopy | 1/585 (0.2%) | 0/585 (0%) | 0/565 (0%) | NA |  | NA |  | NA |  |
| Cord blood parasitemia by qPCR | 19/578 (3.3%) | 6/575 (1.0%) | 4/554 (0.7%) | 0.32 (0.13-0.79) | 0.014 | 0.22 (0.08-0.64) | 0.0056 | 0.69 (0.20-2.44) | 0.57 |
| Parasites detected by histopathology | 27/599 (4.5%) | 6/599 (1.0%) | 3/566 (0.5%) | 0.22 (0.09-0.53) | 0.0008 | 0.12 (0.04-0.39) | 0.0004 | 0.53 (0.13-2.11) | 0.37 |
| High grade pigment detected by histopathology | 27/599 (4.5%) | 1/599 (0.2%) | 5/566 (0.9%) | 0.04 (0.01-0.27) | 0.0012 | 0.20 (0.08-0.51) | 0.0007 | 5.29 (0.62-45.2) | 0.13 |
| Any parasites or pigment detected by histopathology | 316/599 (52.8%) | 179/599 (29.9%) | 226/566 (39.9%) | 0.57 (0.49-0.65) | <0.0001 | 0.76 (0.67-0.86) | <0.0001 | 1.34 (1.14-1.57) | 0.004 |

<sup>1</sup> SP = sulfadoxine-pyrimethamine, DP = dihydroartemisinin-piperaquine
<sup>2</sup> Relative risk for all outcomes with the exception of symptomatic malaria where incidence rate ratio used
<sup>3</sup> Episodes of symptomatic malaria (incidence per person year at risk following initiation of study drugs)
<sup>4</sup> Measured at the time of routine visits conducted every 4 weeks following initiation of study drugs
<sup>5</sup> Hemoglobin measured at 20, 28, and 36 gestational weeks following initiation of study drugs

### Safety and tolerability outcomes

Compliance with study drugs was high, with only 0.5% (81/15568) of routine visits missed when participants were scheduled to receive their first daily dose of study drugs. In addition, only 0.1% (13/15195) and 0.4% (57/15182) of day 2 and day 3 doses of study drugs were reported to have not been taken at home, respectively. Study drugs were well tolerated, with <0.1% of doses associated with vomiting. There were no significant differences in the incidence of any grade 3-4 adverse events, serious adverse events, or grade 3-4 adverse events possibly related to study drugs between the three treatment arms (Table 4). Of note, congenital anomalies occurred in 23 of 1696 (1.4%) deliveries in dihydroartemisinin-piperaquine containing arms compared to 4 of 842 (0.5%) in the sulfadoxine-pyrimethamine arm (p=0.042). Congenital anomalies included 12 episodes of polydactyly, with 11 of 12 in the dihydroartemisinin-piperaquine arms.

**Table 4.** Safety outcomes.

| Outcome | SP<sup>1</sup> | DP<sup>1</sup> | DP+SP<sup>1</sup> | DP vs. SP<sup>1</sup> |  | DP+SP vs. SP<sup>1</sup> |  | DP+SP vs. DP<sup>1</sup> |  |
| --- | --- | --- | --- | --- | --- | --- | --- | --- | --- |
| Prevalence measures | n/N (%) |  |  | RR (95% CI) | P value | RR (95% CI) | P value | RR (95% CI) | P value |
| Post-dosing QTc > 500 ms or interval increase ≥ 60 ms <sup>2</sup> | 3/263 (1.1%) | 4/279 (1.4%) | 4/265 (1.5%) | 1.36 (0.24-7.82) | 0.73 | 1.31 (0.23-7.46) | 0.76 | 1.06 (0.27-4.14) | 0.93 |
| Incidence measures | Events (incidence per person year at risk) |  |  | IRR (95% CI) | P value | IRR (95% CI) | P value | IRR (95% CI) | P value |
| Common adverse events of any severity <sup>3</sup> |  |  |  |  |  |  |  |  |  |
| Abdominal pain | 1532 (3.54) | 1431 (3.30) | 1364 (3.15) | 0.94 (0.85-1.03) | 0.18 | 0.89 (0.81-0.99) | 0.031 | 0.95 (0.86-1.06) | 0.37 |
| Cough | 1064 (2.46) | 1130 (2.61) | 1004 (2.32) | 1.06 (0.95-1.19) | 0.27 | 0.94 (0.84-1.05) | 0.26 | 0.88 (0.79-0.98) | 0.025 |
| Headache | 947 (2.19) | 865 (1.99) | 769 (1.78) | 0.91 (0.82-1.02) | 0.12 | 0.81 (0.72-0.91) | 0.0004 | 0.89 (0.79-1.00) | 0.054 |
| Dysuria | 379 (0.88) | 324 (0.75) | 324 (0.75) | 0.85 (0.72-1.01) | 0.073 | 0.86 (0.72-1.02) | 0.087 | 1.00 (0.84-1.20) | 0.96 |
| Rhinorrhea | 189 (0.44) | 164 (0.38) | 167 (0.39) | 0.87 (0.69-1.09) | 0.22 | 0.88 (0.70-1.10) | 0.27 | 1.02 (0.81-1.28) | 0.88 |
| Diarrhea | 174 (0.40) | 143 (0.33) | 140 (0.32) | 0.82 (0.64-1.05) | 0.12 | 0.80 (0.62-1.04) | 0.097 | 0.98 (0.75-1.27) | 0.87 |
| Fatigue | 101 (0.23) | 79 (0.18) | 56 (0.13) | 0.78 (0.55-1.11) | 0.17 | 0.56 (0.38-0.81) | 0.0020 | 0.71 (0.48-1.07) | 0.10 |
| Chills | 67 (0.15) | 90 (0.21) | 66 (0.15) | 1.32 (0.91-1.92) | 0.14 | 0.97 (0.67-1.42) | 0.89 | 0.73 (0.51-1.05) | 0.089 |
| Epigastric pain | 83 (0.19) | 79 (0.18) | 62 (0.14) | 0.96 (0.68-1.35) | 0.80 | 0.75 (0.53-1.06) | 0.10 | 0.78 (0.53-1.15) | 0.21 |
| Grade 3-4 adverse events <sup>4</sup> |  |  |  |  |  |  |  |  |  |
| Anemia | 53 (0.12) | 28 (0.06) | 44 (0.10) | 0.59 (0.37-0.94) | 0.025 | 0.82 (0.55-1.23) | 0.34 | 1.38 (0.87-2.23) | 0.17 |
| Stillbirth | 15 (0.03) | 16 (0.04) | 7 (0.016) | 1.19 (0.59-2.42) | 0.62 | 0.46 (0.19-1.13) | 0.092 | 0.39 (0.16-0.94) | 0.036 |
| Congenital anomaly | 4 (0.009) | 13 (0.03) | 10 (0.023) | 3.64 (1.19-11.2) | 0.024 | 2.47 (0.78-7.90) | 0.13 | 0.68 (0.30-1.55) | 0.36 |
| Neutropenia | 5 (0.012) | 10 (0.023) | 7 (0.016) | 2.24 (0.77-6.56) | 0.14 | 1.39 (0.44-4.37) | 0.58 | 0.61 (0.24-1.63) | 0.33 |
| Proteinuria | 3 (0.007) | 4 (0.009) | 8 (0.018) | 1.49 (0.33-6.68) | 0.60 | 2.64 (0.70-9.96) | 0.15 | 1.77 (0.53-5.87) | 0.35 |
| Thrombocytopenia | 7 (0.016) | 1 (0.002) | 2 (0.004) | 0.16 (0.02-1.30) | 0.087 | 0.28 (0.06-1.36) | 0.12 | 1.77 (0.16-19.5) | 0.64 |
| Postpartum hemorrhage | 2 (0.005) | 3 (0.007) | 2 (0.004) | 1.68 (0.28-10.1) | 0.57 | 0.99 (0.14-7.04) | 0.99 | 0.59 (0.10-3.53) | 0.56 |
| Pre-eclampsia | 3 (0.007) | 0 (0) | 2 (0.004) | NA |  | 0.66 (0.11-3.95) | 0.65 | NA |  |
| Preterm birth < 28 weeks gestational age | 0 (0) | 0 (0) | 3 (0.007) | NA |  | NA |  | NA |  |
| Elevated temperature | 2 (0.005) | 1 (0.002) | 0 (0) | 0.56 (0.05-6.18) | 0.64 | NA |  | NA |  |
| Any grade 3-4 adverse events | 99 (0.23) | 81 (0.19) | 90 (0.21) | 0.92 (0.68-1.23) | 0.56 | 0.90 (0.68-1.20) | 0.48 | 0.98 (0.73-1.33) | 0.91 |
| Any serious adverse events | 25 (0.06) | 26 (0.06) | 27 (0.06) | 1.17 (0.67-2.02) | 0.58 | 1.07 (0.62-1.84) | 0.81 | 0.92 (0.54-1.57) | 0.76 |
| Grade 3-4 adverse events possible related to study drugs | 50 (0.12) | 29 (0.07) | 41 (0.09) | 0.65 (0.41-1.03) | 0.07 | 0.81 (0.54-1.23) | 0.33 | 1.25 (0.79-2.01) | 0.36 |

<sup>1</sup> SP = sulfadoxine-pyrimethamine, DP = dihydroartemisinin-piperaquine
<sup>2</sup> For the first 300 participants enrolled, electrocardiograms were done at 20, 28, and 36 weeks gestation weeks just before the first dose and 2-6 hours after the third dose of study drugs
<sup>3</sup> Includes only those with at least 200 total events
<sup>4</sup> Includes only those with at least 3 total events

## DISCUSSION

In this double-blind, randomized controlled trial of monthly IPTp, the malaria burden was high in women who received sulfadoxine-pyrimethamine, the current standard-of-care, with malaria parasites detected in nearly half of these participants at the time of monthly routine visits after initiation of IPTp. Dihydroartemisinin-piperaquine markedly reduced the malaria burden, but this did not translate into overall improvements in birth outcomes. While infants born to women who received dihydroartemisinin-piperaquine had a lower risk of preterm birth than those born to women who received sulfadoxine-pyrimethamine, they also had a lower mean birthweight and higher risk of being small-for-gestational age. Combining dihydroartemisinin-piperaquine plus sulfadoxine-pyrimethamine did not improve malaria-related outcomes compared to dihydroartemisinin-piperaquine alone and was associated with a lower mean birthweight and higher risk of small-for-gestational age compared to sulfadoxine-pyrimethamine alone. Thus, although the combination of dihydroartemisinin-piperaquine plus sulfadoxine-pyrimethamine did provide superior antimalarial activity compared to sulfadoxine-pyrimethamine alone, the combination did not result in an improvement in birth outcomes compared to the current standard-of-care.

The finding that IPTp with dihydroartemisinin-piperaquine had far superior antimalarial activity compared to IPTp with sulfadoxine-pyrimethamine is consistent with findings from other studies from eastern and southern Africa, where *P. falciparum* resistance to sulfadoxine-pyrimethamine is widespread [11, 13–16]. In a recent meta-analysis of six trials contributing data on 6646 pregnancies, IPTp with dihydroartemisinin-piperaquine was associated with a 69% lower incidence of symptomatic malaria, 62% lower risk of placental parasitaemia, and 17% lower risk of maternal anemia compared to IPTp with sulfadoxine-pyrimethamine. However, the superior antimalarial activity of dihydroartemisinin-piperaquine did not translate into an improvement in birth outcomes. Indeed, in this meta-analysis, sulfadoxine-pyrimethamine was associated with 34 gm/week higher mean maternal weight gain, a 50 gm higher mean birthweight, and a 15% lower risk of small-for-gestational age compared to dihydroartemisinin-piperaquine [5]. Observational studies also support these results, showing that the use of IPTp with sulfadoxine-pyrimethamine was associated with higher birthweights and a decreased risk of low birthweight in a dose response manner. This effect was observed in areas where antimalarial resistance to sulfadoxine-pyrimethamine was high [17] and in regions with very low malaria transmission, suggesting benefits were independent of the antimalarial activity of sulfadoxine-pyrimethamine [18, 19]. These findings might be explained by non-malarial activities of sulfadoxine-pyrimethamine, which has antibacterial properties, and which may act to improve fetal growth and maternal weight gain. The specific mechanisms of this effect, if present, are unclear, although several studies have demonstrated positive impacts of sulfadoxine-pyrimethamine on the intestinal flora and/or nutrient absorption [2, 6, 20, 21]. and prevention of non-malarial infections [15, 22]. However, in contrast to summary estimates from the meta-analysis, our study, conducted in a region with a very high malaria burden, found that compared to sulfadoxine-pyrimethamine, dihydroartemisinin-piperaquine was associated with a 48% reduction in the risk of preterm birth, which likely can be explained by its potent antimalarial activity, particularly against active placental malaria infection [23].

We hypothesized that IPTp combining the superior antimalarial properties of dihydroartemisinin-piperaquine with the non-malarial activities of sulfadoxine-pyrimethamine would provide superior prevention against adverse birth outcomes compared to either drug used alone.

However, the combination failed to improve birth outcomes and was associated with lower maternal weight gain, lower birthweight, and a higher risk of being born small-for-gestational age compared to sulfadoxine-pyrimethamine alone. These results suggest the possibility that dihydroartemisinin-piperaquine may negatively affect fetal growth, potentially via adversely affecting maternal weight gain. This possibility is supported by a randomized trial in which HIV-infected pregnant women given monthly IPTp with dihydroartemisinin-piperaquine plus daily trimethoprim-sulfamethoxazole (an antifolate similar to sulfadoxine-pyrimethamine) had significantly lower maternal weight gain and non-significantly lower infant birthweight compared to those given daily co-trimoxazole alone [24]. Alternatively, drug-drug interactions may have reduced individual drug exposure, compromising the antimalarial activity of dihydroartemisinin-piperaquine and/or other activities of sulfadoxine-pyrimethamine. This hypothesis is supported by results from a pharmacokinetic study nested in this trial in which women who received the combination had 25% and 34% lower area under the concentration–time curves (AUC) for sulfadoxine and pyrimethamine, respectively, compared to those who received sulfadoxine-pyrimethamine alone and a 19% lower AUC for piperaquine compared to those who received dihydroartemisinin-piperaquine alone [25]. Regardless of the mechanisms involved, the results of this study do not support the use of a combination of dihydroartemisinin-piperaquine plus sulfadoxine-pyrimethamine for IPTp.

As expected, the burden of malaria was much higher in primigravidae than multigravidae [2]. As a result, the absolute benefit of dihydroartemisinin-piperaquine over sulfadoxine-pyrimethamine in reducing the risk of malaria-related outcomes was much greater in primigravidae compared to multigravidae. The risk of adverse birth outcomes was also higher in primigravidae than multigravidae, which may be due to the development of “gravidity dependent immunity”[2]. This likely explains why associations between IPTp regimens and birth outcomes differed by gravidity.

Among primigravidae, who benefitted the most from protection against malaria associated adverse birth outcomes, the net effect of IPTp with dihydroartemisinin-piperaquine was a decreased risk of preterm delivery, but no significant differences in other adverse birth outcomes compared to IPT with sulfadoxine-pyrimethamine. In contrast, the impact of IPTp with dihydroartemisinin-piperaquine on malaria-related birth outcomes was less pronounced among multigravidae, with the net effect being lower birthweight and an increased risk of small-for-gestational age compared to IPTp with sulfadoxine-pyrimethamine. These findings suggest that, with high malaria transmission intensity, IPTp with dihydroartemisinin-piperaquine may offer benefits over sulfadoxine-pyrimethamine in primigravidae, but not in multigravidae.

The results of this study must be considered in light of particularly high-level resistance of *P. falciparum* to sulfadoxine and pyrimethamine in Uganda, mediated by five mutations in PfDHFR (N51I, C59R, and S108N) and PfDHPS (A437G and K540E) that are associated with poor preventive efficacy of sulfadoxine-pyrimethamine and highly prevalent in Uganda [12, 26]. Of concern, two additional mutations that mediate even higher-level resistance, PfDHFR I164L and PfDHPS A581G, are increasing in prevalence in Uganda [12, 27]. Thus, it is not surprising that the malarial preventive efficacy of sulfadoxine-pyrimethamine appears to be very limited in Uganda, and the efficacy of this regimen might be better in regions, such as West Africa, with lower levels of drug resistance.

Tolerability and safety are important considerations when evaluating drugs for routine use during pregnancy. All three IPTp regimens were well tolerated, with no significant differences in the incidence of any grade 3-4 adverse events or serious adverse events. Of concern, 11 of 12 infants born with polydactyly had mothers who received dihydroartemisinin-piperaquine. These findings are similar to those from a previous study by our group from the same study area comparing monthly IPTp with dihydroartemisinin-piperaquine versus sulfadoxine-pyrimethamine, in which 9 of 10 infants born with polydactyly had mothers who received dihydroartemisinin-piperaquine [11]. Polydactyly has been amongst the most commonly observed congenital anomalies in African populations, and genetic factors have been postulated to explain this high frequency [28, 29]. A causal link between IPTp with dihydroartemisinin-piperaquine and polydactyly appears unlikely from an embryologic standpoint, given limb development occurs in the first trimester, before initiation of IPTp [30]. In addition, an increased risk of congenital anomalies or polydactyly has not been reported in other studies of IPTp with dihydroartemisinin-piperaquine [13–16]. However, a significantly increased risk of polydactyly in the dihydroartemisinin-piperaquine arms of two independent trials of IPTp offers some reason for caution when considering potential benefits of this regimen.

This study had some limitations. It was conducted in an area of high transmission intensity with widespread resistance to sulfadoxine-pyrimethamine, limiting generalizability to other settings. Only the first daily dose of study drugs was directly observed, which could have differentially affected exposure to dihydroartemisinin-piperaquine, a three-day regimen, although reported compliance to the second and third doses administered at home was high. P values were not adjusted for multiple comparisons, warranting cautious interpretation. The study was not powered to detect differences in individual components of the primary endpoint or between gravidity subgroups. Thus, non-statistically significant associations should not necessarily be interpreted as an absence of effect. Lastly, findings presented in this report did not include investigations of potential mechanisms by which IPTp regimens may have affected birth outcomes independent of their antimalarial activity.

This study provides further evidence that, in areas with high level *P. falciparum* resistance to sulfadoxine-pyrimethamine, the burden of malaria may be unacceptably high among pregnant women administered the current standard of care for IPTp. Replacing sulfadoxine-pyrimethamine with dihydroartemisinin-piperaquine for IPTp would likely result in significant reductions in clinical malaria and maternal anemia, especially among primigravidae. However, such a change may negatively impact fetal growth. Combining dihydroartemisinin-piperaquine and sulfadoxine-pyrimethamine offered promise, but it provided no clear benefits, raising the possibility that dihydroartemisinin-piperaquine adversely affects maternal weight gain and fetal growth.

Furthermore, co-administration of these drugs negatively affected individual drug exposure. Further studies are needed to better elucidate the mechanisms by which dihydroartemisinin-piperaquine and sulfadoxine-pyrimethamine affect fetal weight gain independent of antimalarial activity. Research is also needed to identify new interventions, such as alternative IPTp regimens, vaccines, or monoclonal antibodies to better prevent malaria in pregnancy, reduce the risk of adverse birth outcomes, and ultimately improve the health of pregnant women and their infants.

## Data Availability

All data used in the analyses are available on the ClinEpiDB website, https://clinepidb.org/ce/app

## ACKNOWLEDGEMENTS

We thank the women for their participation in the study; Dr. Wabwire Mathias Panyako, the district health officer of Busia district, Uganda, and the dedicated staff of Masafu General Hospital for their support and hosting the study; and the members of the Tororo-Busia community advisory board for supporting community engagement. We are also grateful to Thomas Katairo for generating data on drug resistance markers and the staff of the Infectious Diseases Research Collaboration and the University of California, San Francisco, for their administrative and regulatory support.

## AUTHOR CONTRIBUTIONS

AK, PJR, MRK, MER, and GD conceived the idea for the study. AK, JIN, PK, and GD wrote the protocol. JK, MA, HA, and MN were responsible for management of the study participants. HA, JA, PO, NO, EC, SG, and PJR contributed to laboratory studies. GD and MER performed the statistical analysis with inputs from AK. AK and GD wrote the first draft of the manuscript. All authors interpreted the data and critically reviewed the manuscript.

## SUPPORTING INFORMATION

S1 CONSORT Checklist. CONSORT checklist (PDF)

S2 Protocol. Study protocol (PDF)

S3 Analysis. Statistical analysis plan (PDF)

## Notes

### Competing Interest Statement

The authors have declared no competing interest.

### Clinical Trial

The study was registered with ClinicalTrials.gov (NCT04336189 https://clinicaltrials.gov/)

### Funding Statement

Yes

### Author Declarations

The study was approved by the Makerere University School of Biomedical Sciences Research Ethics Committee (SBS 714), the Uganda National Council for Science and Technology (HS 2746), the Uganda National Drug Authority (CTC 0135/2020), and the University of California San Francisco Human Research Protection Program (19-29105).

## REFERENCES

1. World Health Organization. World Malaria Report 2024: addressing inequity in the global malaria response. Geneva, Switzerland: World Health Organization; 2024.

2. Desai M, ter Kuile FO, Nosten F, McGready R, Asamoa K, Brabin B, Newman RD. Epidemiology and burden of malaria in pregnancy. Lancet Infect Dis. 2007;7(2):93-104.

3. World Health Organization. WHO guidelines for malaria, 3 June 2022. World Health Organization, 2022.

4. Flegg JA, Humphreys GS, Montanez B, Strickland T, Jacome-Meza ZJ, Barnes KI, et al. Spatiotemporal spread of *Plasmodium falciparum* mutations for resistance to sulfadoxine-pyrimethamine across Africa, 1990-2020. PLoS Comput Biol. 2022;18(8):e1010317.

5. Roh ME, Gutman J, Murphy M, Hill J, Madanitsa M, Kakuru A, et al. Dihydroartemisinin-piperaquine versus sulfadoxine-pyrimethamine for intermittent preventive treatment of malaria in pregnancy: a systematic review and individual participant data meta-analysis. medRxiv. 2024. Epub 20241126. doi: 10.1101/2024.11.23.24315401.

6. Roh ME, Kuile FOT, Rerolle F, Glymour MM, Shiboski S, Gosling R, et al. Overall, anti-malarial, and non-malarial effect of intermittent preventive treatment during pregnancy with sulfadoxine-pyrimethamine on birthweight: a mediation analysis. Lancet Glob Health. 2020;8(7):e942–e53.

7. Division of AIDS (DAIDS) Table for grading the severity of adult and pediatric adverse events, version 2.0. Washington DC: US. Department of Health and Human Services, National Institutes of Health, Naitional Institute of Allergy and Infectious Diseases, Division of AIDS, 2014. http://www.niaid.nih.gov/LabsAndResources/resources/DAIDSClinRsrch/Documents/daidsaegradingtable.pdf.

8. Hofmann N, Mwingira F, Shekalaghe S, Robinson LJ, Mueller I, Felger I. Ultra-sensitive detection of *Plasmodium falciparum* by amplification of multi-copy subtelomeric targets. PLoS Med. 2015;12(3):e1001788.

9. Villar J, Cheikh Ismail L, Victora CG, Ohuma EO, Bertino E, Altman DG, et al. International standards for newborn weight, length, and head circumference by gestational age and sex: the Newborn Cross-Sectional Study of the INTERGROWTH-21st Project. Lancet. 2014;384(9946):857-68.

10. Ategeka J, Kakuru A, Kajubi R, Wasswa R, Ochokoru H, Arinaitwe E, et al. Relationships between measures of malaria at delivery and adverse birth outcomes in a high-transmission area of Uganda. J Infect Dis. 2020;222(5):863–70.

11. Kajubi R, Ochieng T, Kakuru A, Jagannathan P, Nakalembe M, Ruel T, et al. Monthly sulfadoxine-pyrimethamine versus dihydroartemisinin-piperaquine for intermittent preventive treatment of malaria in pregnancy: a randomized controlled trial. The Lancet 2019; 393(10179):1428–1439.

12. Asua V, Conrad MD, Aydemir O, Duvalsaint M, Legac J, Duarte E, et al. Changing prevalence of potential mediators of aminoquinoline, antifolate, and artemisinin resistance across Uganda. J Infect Dis. 2021;223(6):985–94.

13. Desai M, Gutman J, L’Lanziva A, Otieno K, Juma E, Kariuki S, et al. Intermittent screening and treatment or intermittent preventive treatment with dihydroartemisinin-piperaquine versus intermittent preventive treatment with sulfadoxine-pyrimethamine for the control of malaria during pregnancy in western Kenya: an open-label, three-group, randomised controlled superiority trial. Lancet. 2015;386(10012):2507–19.

14. Kakuru A, Jagannathan P, Muhindo MK, Natureeba P, Awori P, Nakalembe M, et al. Dihydroartemisinin-piperaquine for the prevention of malaria in pregnancy. N Engl J Med. 2016;374(10):928–39.

15. Madanitsa M, Barsosio HC, Minja DTR, Mtove G, Kavishe RA, Dodd J, et al. Effect of monthly intermittent preventive treatment with dihydroartemisinin-piperaquine with and without azithromycin versus monthly sulfadoxine-pyrimethamine on adverse pregnancy outcomes in Africa: a double-blind randomised, partly placebo-controlled trial. Lancet. 2023;401(10381):1020–36.

16. Mlugu EM, Minzi O, Kamuhabwa AAR, Aklillu E. Effectiveness of intermittent preventive treatment with dihydroartemisinin-piperaqunine against malaria in pregnancy in Tanzania: a randomized controlled trial. Clin Pharmacol Ther. 2021;110(6):1478–89.

17. Desai M, Gutman J, Taylor SM, Wiegand RE, Khairallah C, Kayentao K, et al. Impact of sulfadoxine-pyrimethamine resistance on effectiveness of intermittent preventive therapy for malaria in pregnancy at clearing infections and preventing low birth weight. Clin Infect Dis. 2016;62(3):323–33.

18. Cellich P, Unger HW, Rogerson SJ, Mola GDL. Impact on pregnancy outcomes of intermittent preventive treatment with sulfadoxine-pyrimethamine in urban and peri-urban Papua New Guinea: a retrospective cohort study. Malar J. 2024;23(1):201.

19. Chico RM, Chaponda EB, Ariti C, Chandramohan D. Sulfadoxine-pyrimethamine exhibits dose- response protection against adverse birth outcomes related to malaria and sexually transmitted and reproductive tract infections. Clin Infect Dis. 2017;64(8):1043–51.

20. Kim S, Naziripour A, Prabhala P, Horváth V, Junaid A, Breault DT, et al. Direct therapeutic effect of sulfadoxine-pyrimethamine on nutritional deficiency-induced enteric dysfunction in a human Intestine Chip. EBioMedicine. 2024;99:104921.

21. Waltmann A, McQuade ETR, Chinkhumba J, Operario DJ, Mzembe E, Itoh M, et al. The positive effect of malaria IPTp-SP on birthweight is mediated by gestational weight gain but modifiable by maternal carriage of enteric pathogens. EBioMedicine. 2022;77:103871.

22. Lee JJ, Kakuru A, Jacobson KB, Kamya MR, Kajubi R, Ranjit A, et al. Monthly sulfadoxine- pyrimethamine during pregnancy prevents febrile respiratory illnesses: a secondary analysis of a malaria chemoprevention trial in Uganda. Open Forum Infect Dis. 2024;11(4):ofae143.

23. Rogerson SJ, Hviid L, Duffy PE, Leke RF, Taylor DW. Malaria in pregnancy: pathogenesis and immunity. Lancet Infect Dis 2007; 7(2): 105–17.

24. Barsosio HC, Madanitsa M, Ondieki ED, Dodd J, Onyango ED, Otieno K, et al. Chemoprevention for malaria with monthly intermittent preventive treatment with dihydroartemisinin-piperaquine in pregnant women living with HIV on daily co-trimoxazole in Kenya and Malawi: a randomised, double-blind, placebo-controlled trial. Lancet. 2024;403(10424):365–78.

25. Mwebaza N, Roh ME, Geng YZ, Opio L, Opira B, Marzan F, et al. Drug-Drug Interaction Between Dihydroartemisinin-Piperaquine and Sulfadoxine-Pyrimethamine During Malaria Chemoprevention in Pregnant Women. Clinical Pharmacol Ther 2025;117(2):506–514.

26. Nankabirwa JI, Wandera B, Amuge P, Kiwanuka N, Dorsey G, Rosenthal PJ, et al. Impact of intermittent preventive treatment with dihydroartemisinin-piperaquine on malaria in Ugandan schoolchildren: a randomized, placebo-controlled trial. Clin Infect Dis. 2014;58(10):1404–12.

27. Kreutzfeld O, Tumwebaze PK, Byaruhanga O, Katairo T, Okitwi M, Orena S, et al. Decreased Susceptibility to Dihydrofolate Reductase Inhibitors Associated With Genetic Polymorphisms in Ugandan Plasmodium falciparum Isolates. J Infect Dis. 2022;225(4):696–704.

28. Scott-Emuakpor AB, Madueke ED. The study of genetic variation in Nigeria. II. The genetics of polydactyly. Hum Hered. 1976;26(3):198-202.

29. Sevene E, Bardají A, Mariano A, Machevo S, Ayala E, Sigaúque B, et al. Drug exposure and pregnancy outcome in Mozambique. Paediatr Drugs. 2012;14(1):43–9.

30. Rogers BH, Schmieg SL, Pehnke ME, Shah AS. Evaluation and Management of Preaxial Polydactyly. Curr Rev Musculoskelet Med. 2020;13(4):545–51.

